# Altered Cytokine Profile in Clinically Suspected Seronegative Autoimmune Associated Epilepsy

**DOI:** 10.1101/2024.09.13.24310337

**Authors:** Katherine Motovilov, Cole Maguire, Deborah Briggs, Esther Melamed

**Affiliations:** The University of Texas at Austin, Department of Neurology

## Abstract

**Background and Objectives:** Autoimmune-associated epilepsy (AAE), a condition which responds favorably to immune therapies but not traditional anti-seizure interventions, is emerging as a significant contributor to cases of drug-resistant epilepsy. Current standards for the diagnosis of AAE rely on screening for known neuronal autoantibodies in patient serum or cerebrospinal fluid. However, this diagnostic method fails to capture a subset of drug-resistant epilepsy patients with suspected AAE who respond to immunotherapy yet remain seronegative (snAAE) for known autoantibodies.

**Methods:** To identify potential biomarkers for snAAE, we evaluated the most comprehensive panel of assayed cytokines and autoantibodies to date, comparing patients with snAAE, anti-seizure medication (ASM) responsive epilepsy, and patients with other neuroinflammatory diseases.

**Results:** We found a unique signature of 14 cytokines significantly elevated in snAAE patients including: GM-CSF, MCP-2/CCL8, MIP-1a/CCL3, IL-1RA, IL-6, IL-8, IL-9, IL-10, IL-15, IL-20, VEGF-A, TNF-b, LIF, and TSLP. Based on prior literature, we highlight IL-6, IL-8, IL-10, IL-13, VEGF-A, and TNF-b as potentially actionable cytokine biomarkers for snAAE, which could be of diagnostic utility in clinical evaluations of snAAE patients. Autoantibody-ome screening failed to identify autoantibodies targeting neuronal channel proteins in snAAE patients. Interestingly, ASM-responsive epilepsy patients displayed elevations in the proportion of autoantibodies targeting brain plasma membrane proteins, possibly pointing to the presence of immune hyperactivity/dysfunction despite well-controlled seizure activity and suggesting ASM-responsive patients may experience disease progression independent of seizure activity (PISA).

**Discussion:** Overall, our findings suggest that simply expanding existing autoantibody screens may not sufficiently enhance diagnostic power for snAAE. Instead, we propose that cytokine analysis may serve as a promising diagnostic avenue for identifying immune dysregulation in AAE patients and enabling opportunities for trials of immunotherapies.

## Introduction

Approximately 20-40% of individuals with epilepsy have drug-resistant epilepsy (DRE) characterized by resistance to multiple anti-seizure medications (ASMs)^1^. Of these DRE patients, at least 20% are thought to be autoimmune-associated epilepsies (AAE), characterized by a favorable response to immunotherapies such as high dose steroids, intravenous immunoglobulin (IVIG), and plasma exchange (PLEX)^2^. Currently, the standard for diagnosing AAE is the identification of autoantibodies to known neuronal autoantigens (e.g. GAD65, NMDAR, LGI1, VGK, etc)^2^. As a result, most AAE studies have focused on seropositive autoimmune-associated epilepsy (spAAE) patients.

In addition to spAAE, there is also a subset of DRE patients who fail surgery and/or implantation of epileptic devices. These DRE patients may present with a *chronic*, super refractory seizure profile similar to spAAE and respond to immunosuppressive therapies, though remain seronegative to known neuronal autoantigens. Despite the lack of a known neuronal autoantibodies, the immune system is still thought to play a critical role in the pathology of these seronegative patients who are often clinically suspected of having seronegative autoimmune epilepsy (snAAE)^3^.

Compared to spAAE, snAAE has been more challenging to diagnose and treat due to the lack of clinically available biomarkers and accepted diagnostic framework. Diagnosis is further complicated by ongoing discussion in the field regarding whether the etiology of snAAE is autoimmune (the presence of autoreactivity against a specific autoantigen) or autoinflammatory (characterized by generalized inflammation with an undefined target)^4^. snAAE is typically suspected in individuals with different seizure types, multifocal epileptiform discharges, multi-ASM resistance, and response to immunotherapies, features which are typically captured by high Antibody Prevalence in Epilepsy and Encephalopathy 2 (APE2) and Response to Immunotherapy in Epilepsy and Encephalopathy (RITE^2^) scores^5,6^. Examples of well characterized acute snAAE adjacent syndromes include new-onset refractory status epilepticus (NORSE), febrile infection–related epilepsy syndrome (FIRES), devastating epileptic encephalopathy in school-aged children (DESC), and Rasmussen encephalitis (RE)^7–9^. Although snAAE patients can experience a favorable response to immunotherapies, it remains challenging to obtain payer approval for these treatments due to a lack of documented autoantibodies, which is typically required by insurance companies for AAE diagnosis. Thus, there is a critical need to identify new clinical biomarkers, which can augment autoantibody screening in suspected AAE, as well as improve time to diagnosis and treatment of snAAE patients.

In recent literature, cytokines, including IL-8, IL-12, TNF-B, VEGF-A, and IL-6^10,11^ have been documented to be elevated in snAAE patients, suggesting evidence for ongoing systemic inflammation. To better characterize cytokine abnormalities in chronic refractory snAAE patients, we performed one of the largest cytokine multiplexing studies (panel of 65 cytokines) in snAAE patients, and defined systemic cytokine signatures elevated in this population. In addition, to expand the understanding of involved autoantibodies in epilepsy patients, we conducted a comprehensive screen of ∼24,000 autoantibodies to identify potentially putative neuronal autoantigens.

Our findings expand on the understanding of AAE pathophysiology, highlighting the role of cytokine and autoantibody dysregulation in AAE patients. In addition, our results suggest that incorporation of cytokines alongside autoantibodies could improve diagnostic criteria for snAAE and offer opportunities for the use of targeted immune therapies.

## Methods

### Standard Protocol Approvals, Registrations, and Patient Consents

Patients with known neurological conditions including multiple sclerosis (MS), ASM-responsive epilepsy, snAAE, as well as healthy controls were enrolled between November 11^th^, 2019 and March 20^th^, 2020 at the Dell Medical School Neuroimmunology Center at The University of Texas at Austin and Dell Seton Medical Center. Patient data were anonymized for analysis. The study was approved by the UT Austin Institutional Review Board (IRB ID: 2016070072) and was in compliance with the October 2013 Declaration of Helsinki principles. All subjects gave written informed consent in adherence to local and national regulations.

### Defining Clinically Suspected Seronegative Autoimmune-Associated Epilepsy and Non-Autoimmune-Associated Epilepsy

snAAE patients were selected from refractory epilepsy patients who responded to immunomodulatory therapy, including corticosteroids or IVIG, tested seronegative for known neuronal antibodies, and had an APE2 score greater than 4^6^. Non-autoimmune epilepsy patients were characterized by hospitalized epilepsy patients who responded well to ASMs, (ie resolution of seizures) and whose APE2 score was less than 4.

### Human Sample Collection and Serum Processing

Venous blood was collected in Greiner Bio-One Serum Clot Activating Tubes (Greiner Bio-One 455010P) and stored at room temperature. Blood samples for snAAE patients were collected prior to corticosteroids or IVIG administration. The samples were then spun at 2000 g for 10 minutes at 4°C, before aliquoting the sera and storing at -80°C.

### Cytokine Multiplexing

65 cytokines (Table 1) were measured in patient serum using Eve Technologies’ HD65 Cytokine Multiplexing assay, of which IL3 and IL4 for the PCA analysis were dropped due to respectively: not being within the limits of detection in any sample and a missing value in an snAAE sample. Differences for individual cytokines between groups were evaluated by a Wilcoxon rank sum test with Benjamini Hochberg corrections, and Principal Component Analysis (PCA) was used to visualize sample variation between groups.

**Table 1.** Measured Cytokines.

| **65 Cytokines Analyzed** |  |  |
| --- | --- | --- |
| EGF | IL-17A | BCA-1 |
| FGF-2 | IL-1RA | MCP-4 |
| Eotaxin | IL-1a | I-309 |
| TGF-a | IL-9 | IL-16 |
| G-CSF | IL-1B | TARC |
| Flt-3L | IL-2 | X6Ckine |
| GM-CSF | IL-5 | Eotaxin-3 |
| Fractalkine | IL-6 | LIF |
| IFNa2 | IL-7 | TPO |
| IFNy | IL-8 | SCF |
| GRO-alpha | IP-10 | TSLP |
| IL-10 | MCP-1 | IL-33 |
| MCP-3 | MIP-1a | IL-20 |
| IL-12P40 | MIP-1b | IL-21 |
| MDC-30 | RANTES | IL-23 |
| IL-12P70 | TNFa | TRAIL |
| PDGF-AA | TNFb | CTACK |
| IL-13 | VEGF-A | SDF-1a-B |
| PDGF-BB | IL-18 | ENA-78 |
| IL-15 | Eotaxin-2 | MIP-1d |
| sCD40L | MCP-2 | IL-28A |
| IL-4 | IL-5 |  |

### Phage Viral Display Autoantibody Sequencing (PhIP-seq)

Human serum samples were evaluated for autoantibodies using the CDI Labs Human Proteome Phage-Display (HuScan) to identify reactive antibodies against human linear protein epitopes. Reads were processed as previously reported with autoantibody “hits” identified using the edgeR package to detect targets that had at least 15 reads, a p-value of less than 0.001, and a fold-change of 5 compared to buffer mock immunoprecipitations as previously reported^12^.

Neurotransmitter-association was defined as all genes in the gene ontologies GO:0007269 and GO:0042136, in addition to genes from any children ontologies under these two sets. Anatomical or cellular location of the protein’s expression (in the central nervous system and on the cell membrane) was retrieved from the Human Protein Atlas^13^. Finally, a list of all ion channel genes was retrieved from HUGO to identify any ion channels with identifiable autoantibodies^14^.

### Data Availability

Data and code are deposited on github at the following link https://github.com/melamedlab/AutoimmuneEpilepsy.

## Results

### Patient Demographics

Our study cohort consisted of thirty-two participants with snAAE (n=5), ASM-responsive epilepsy (n=5), MS (n=15), as well as healthy controls (n=7). All snAAE participants were classified as having focally unaware seizures with some culminating in Generalized Tonic Clonic seizures (GTCs) (Table 2). Participants’ ages ranged from 18-80 years old.

**Table 2.** Demographics of Cohort.

| Group | Sex | Seizure Type | MRI Findings | List of Known ASMs |
| --- | --- | --- | --- | --- |
| snAAE | M | Focal unaware | No MTS - R temporal resection and L frontal trauma | Xcorpi, CZP, Epidiolex, LAC, BRIV |
| snAAE | F | Initially presented with NORSE, focal unaware to GTC | L transverse, sigmoid, and superior internal jugular vein thrombosis; normal parenchyma | VPA, LEV, LAC, CLB, Lorazepam, FYC |
| snAAE | F | Focal unaware to GTC | Normal | LEV |
| snAAE | F | Focal unaware | No MTS; left temporal edema | LEV |
| snAAE | F | Focal epilepsy | Laser ablation of L anterior insula and mesial frontal regions | LEV, LCM, Phenobarbital, CLB, ZNS, Pentobarbital, Ketamine, LZP, DEX |
| ASM-Responsive Epilepsy | M | Epilepsy and PNEE | Normal | PHT |
| ASM-Responsive Epilepsy | M | Focal unaware to GTC | Chiari malformation | ZNS |
| ASM-Responsive Epilepsy | F | Focal unaware to GTC | Small T2 change in corpus callosum | LEV |
| ASM-Responsive Epilepsy | M | Focal to GTC suspected | B cerebellar and L occipital chronic ischemia | LEV |
| ASM-Responsive Epilepsy | F | Focal unaware | No MTS but L and inferior temporal lobe metastasis and prior strokes | LEV |

GTC = general tonic clonic seizure; PNEE = psychogenic non-epileptic events; R = right; L = left; B = both; MTS = mesial temporal sclerosis; T2 = transverse relaxation time; LEV = Levetiracetam; CZP = clonazepam; LCM = Lacosamide; ZNS = Zonisamide; LZP = Lorazepam; DEX = Dexamethasone; VPA = valproic acid; PHT = Phenytoin; LAC = Lacosamide; CLB = clobazam; FYC = Fycompa; BRIV = Brivaracetam; NORSE = New Onset Refractory Status Epilepticus

### Cytokine multiplexing revealed an altered cytokine profile in snAAE patients

We evaluated 65 circulating serum cytokines in a total of 32 patients, with groups comprised of snAAE (n=5), controls (n=7), ASM-responsive epilepsy (n=5), and MS (n=15). Principle coordinate analysis (PCA) of the 63 detected cytokines (Fig 1A) revealed a widely variable and altered cytokine profile in snAAE patients, who clustered separate from healthy controls, ASM-responsive epilepsy, and MS patients. We identified 14 cytokines which explained the observed differences between snAAE and other groups, including granulocyte macrophage colony-stimulating factor (GM-CSF), monocyte chemotactic protein (MCP)-2/chemokine (C-C motif) ligand (CCL)-8, macrophage inflammatory protein-1 alpha (MIP-1a)/CCL3), interleukin (IL)-1RA, IL-6, IL-8, IL-9, IL-10, IL-15, IL-20, vascular endothelial growth factor A (VEGF-A), tumor necrosis factor beta (TNF-b), leukemia inhibitory factor (LIF), thymic stromal lymphopoietin (TSLP) (Fig 1C-P, Supplemental Table 1), all of which were significantly upregulated in the snAAE patients compared to other groups. Additionally, we saw notable increases in epidermal growth factor (EGF), MCP-3, IL-1A, IL-13, IL-16, and IL-28A within snAAE patients (Fig 1Q-V). Although these differences did not attain significance, they were high contributors to the snAAE patient cytokine signature (Fig 1B). This widespread activation of cytokines has been described in other inflammatory conditions such as acute infection and autoimmune disease and is often referred to as a “cytokine storm” ^15^. However, compared to “cytokine storms” seen in other conditions, the cytokines elevated in snAAE patients did not include the potently inflammatory cytokines CXCL10 (IP-10) and tumor necrosis factor alpha (TNF-a) (Supplemental Fig 1). Of note, although data was only available for one of our snAAE patients, we found that all cytokines returned to healthy-control levels post steroid administration (Fig 1C-V).

**Figure 1.**
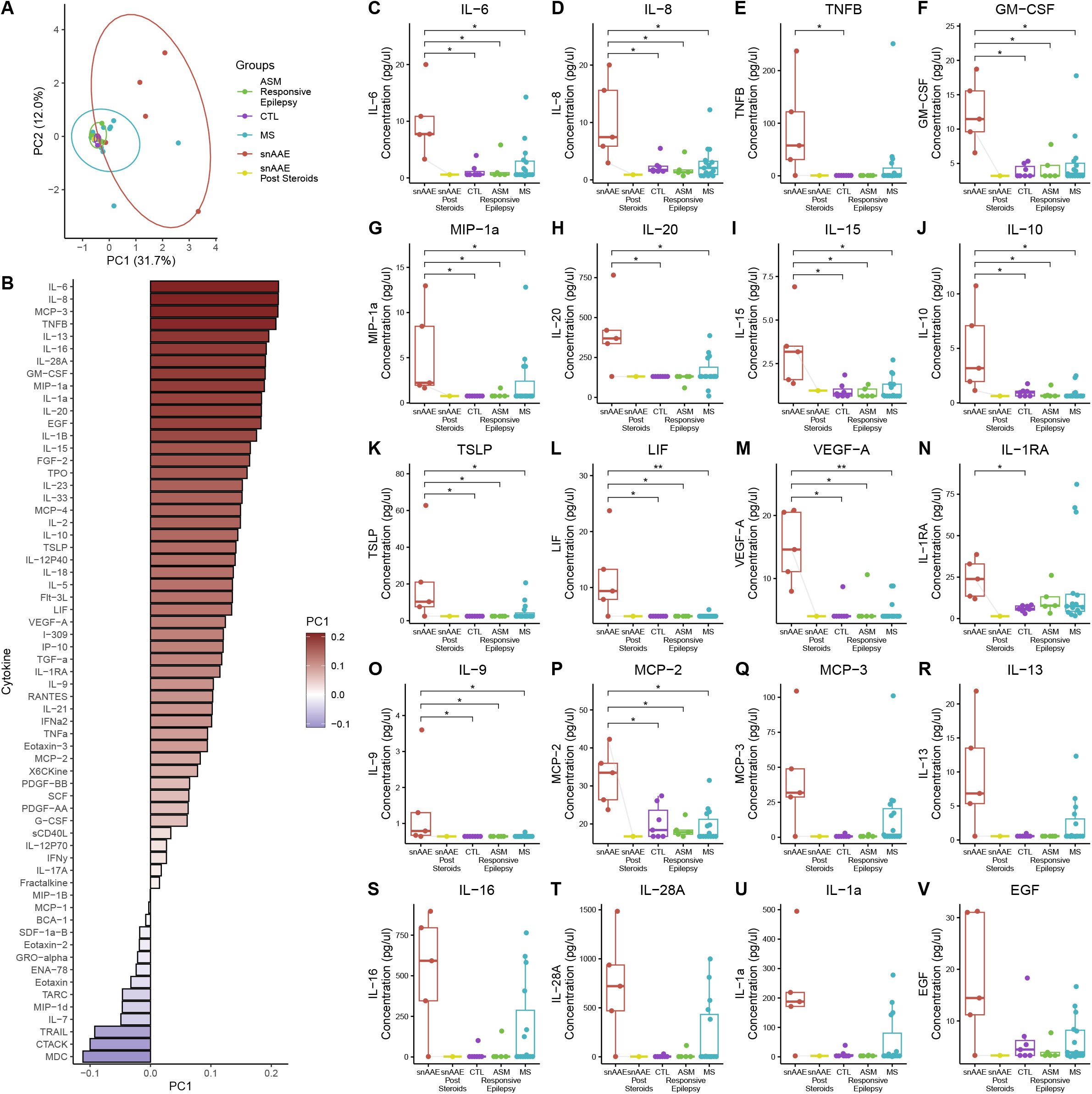
Elevated circulating cytokines in clinically suspected seronegative autoimmune epilepsy patients. **A)** PCA of 63 cytokines separates snAAE patients from other evaluated patients. **B)** Contribution of each cytokine to PC1 from (A). Comparison across snAAE, healthy controls (CTL), ASM-responsive epilepsy, and multiple sclerosis (MS) patients for **C)** IL-6, **D)** IL-8, **E)** TNFB, **F)** GM-CSF, **G)** MIP-1a, **H)** IL-20, **I)** IL-15, **J)** IL-10, **(K)** TSLP, **L)** LIF, **M)** VEGF-A, **N)** IL-1RA, **O)** IL-9, **P)** MCP-2, **Q)** MCP-3, **R)** IL-13, **S)** IL-16, **T)** IL-28A, **U)** IL-1a, **V)** EGF. For one snAAE participant, cytokines were additionally measured post steroid administration, shown in (C-V) connecting their data points before and after steroids with a gray line.

### Lack of Neuronal Autoantibodies in Clinically Suspected Seronegative Autoimmune Epilepsy

To characterize the variation in autoantibody presentation in AAE and probe for novel autoantigens, we evaluated a total of 32 patients who were either snAAE (n=7), ASM-responsive (n=5), healthy controls (n=10), or MS (n=10) for autoantibodies, using the HuScan autoantibody PhIP-seq panel, which screens for autoantibodies to ∼24,000 human proteins. From the 7 snAAE patients, we identified 162 autoantibodies that were not found in the healthy control samples.

To narrow the list to putative biomarker auto-antibodies in snAAE, we evaluated how many patients had autoantibodies that target either i) known ion channels, ii) proteins involved in neurotransmitter production and release, or iii) central nervous system (CNS) cell membrane proteins. Notably, only three snAAE patients had autoantibodies against ion channels: anoctamin 8 (ANO8), calcium voltage-gated channel auxiliary subunit gamma 6 (CACNG6), or calcium voltage-gated channel subunit alpha1 S (CACNA1S) (Fig 2). However, all three of these ion channels have low human brain regional specificity and are primarily expressed in skeletal muscle, making these autoantibodies potentially less likely to be clinically meaningful snAAE candidates^13^. There were also few positive autoantibody hits in snAAE individuals for known proteins involved in neurotransmitter release/production or cell membrane expressed proteins, specifically: calcium dependent sectarian activator (CADPS), ELKS/RAB6-Interacting/CAST Family Member 2 (ERC2), neurofibromin 1 (NF1), neurexin 2 (NRXN2), neurexin 3 (NRXN3), piccolo presynaptic cytomatrix protein (PCLO), syntaxin binding protein 5 (STXBP5), unc-13 homolog B (UNC13B), and unc-13 homolog C (UNC13C) (Fig 2A).

**Figure 2.**
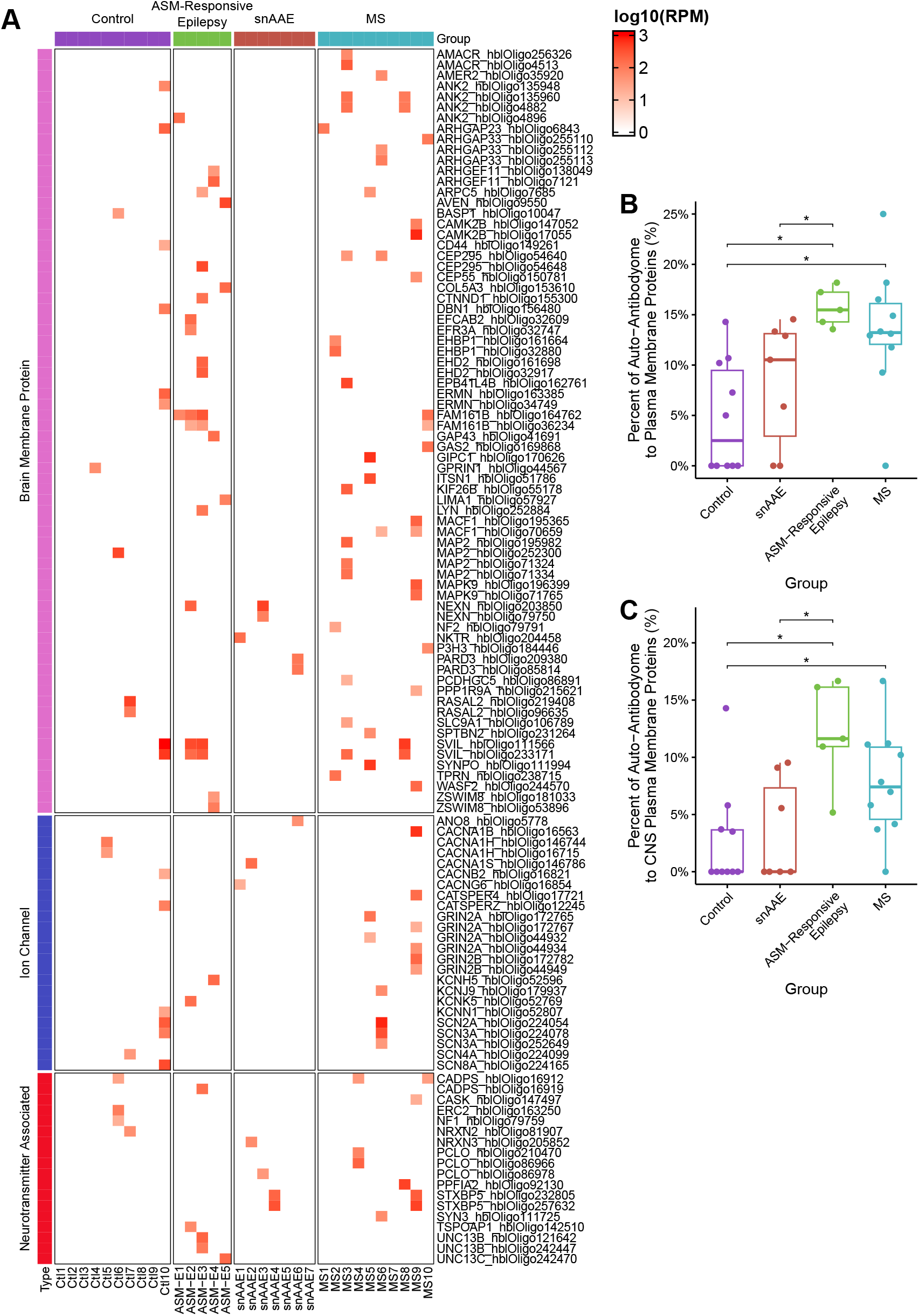
Autoantibody screen identifies ion channel protein autoantibodies in snAAE patients. **A)** Heatmap of significant neurotransmitter, brain plasma membrane, and ion channel autoantigens, where red indicates detection and white indicates no detection/non-significant autoantibody signal. Oligo IDs correspond to the HuScan PhIP-seq platform. Ion channel status was retrieved from the HUGO Gene Nomenclature Committee^14^ and additional brain plasma membrane proteins were identified using the Human Protein Atlas proteinatlas.org^13^. Percent of identified autoantibodies to **B)** proteins expressed on the plasma membrane, and **C)** proteins found on both the plasma membrane and in the CNS.

Interestingly, we also identified that ASM-responsive epilepsy patients had a higher proportion of their autoantibody-ome targeting known plasma membrane proteins (Fig 2B), including those expressed in the CNS (Fig 2C). This elevation could imply that even in well-managed ASM-responsive epilepsy, ongoing seizures and associated inflammation may result in novel auto-reactivity, possibly contributing to disease progression and potentially neurodegeneration.

## Discussion

Autoimmune-associated epilepsy (AAE) has emerged as an important etiology of intractable seizures, presenting a new and exciting avenue for the use of immunomodulatory therapies in patients with refractory epilepsy. Yet, diagnosing AAE remains a challenge due to the scarcity of precise biomarkers for snAAE and the limited number of known autoantibodies in spAAE. Our study leveraged comprehensive cytokine multiplexing and autoantibody-ome screening to evaluate snAAE patients, revealing a distinct cytokine signature. These findings not only deepen our understanding of AAE, but also demonstrate the limitations of current AAE diagnostic framework in capturing snAAE.

We identified 14 cytokines significantly elevated in snAAE individuals compared to individuals with non-immune mediated epilepsies, patients with MS, and healthy controls. Elevated cytokines included: GM-CSF, MCP-2/CCL8, MIP-1a/CCL3, IL-1RA, IL-6, IL-8, IL-9, IL-10, IL-15, IL-20, VEGF-A, TNF-b, LIF, and TSLP. Further, we found notable increases in EGF, MCP-3/CCL7, IL-1A, IL-13, IL-16, and IL-28A, which contribute to the unique cytokine signature of snAAE patients (Fig 1B). Broadly, we found these results corroborate findings from prior studies (Table 3). Elevations in IL-10, which we observed in snAAE patient sera, have been previously demonstrated in multiple other instances, including in the plasma of glutamic acid decarboxylase antibodies positive AAE patients and both the serum and CSF of ASM-resistant patients^16,17^. Further, our observed increases in the potently inflammatory cytokine IL-6 have also previously been described within both suspected AAE patients and patients with NORSE^11,18^. Notably, NORSE patients, specifically, have been previously noted to have many of the other serum cytokine perturbations found in our study, including increases in MIP-1a/CCL3, IL-8, and IL-10, suggesting that some of these cytokine changes may occur in both acute and chronic disease courses^18^. Finally, elevations in IL-8, IL-13, VEGF-A, and TNF-b have also been previously reported in refractory epilepsy patients, a finding which we have now extended specifically to snAAE patients^19^.

**Table 3.** Literature review of cytokine perturbations in AAE and snAAE patients.

| Cytokine | *Our Study* | *Gillinder et al 2021* |  | *Basnyat et al 2023* | *Hanin et al 2023* |  | *Han Y et al 2020* |
| --- | --- | --- | --- | --- | --- | --- | --- |
|  | Serum | Serum | CSF | Plasma | CSF | Serum | CSF |
| GM-CSF | ↑↑ | n.s. | n.s. | - | n.s. | n.s. | - |
| MCP-2 (CCL8) | ↑↑ | - | - | - | - | - | - |
| MIP-1a (CCL3) | ↑↑ | n.s. | n.s. | - | ↑↑ in NORSE | ↑↑ in NORSE | - |
| IL-1RA | ↑↑ | n.s. | n.s. | - | - | - | - |
| IL-6 | ↑↑ | n.s. | n.s. | n.s. | ↑↑ in NORSE | ↑↑ in NORSE | ↑↑ in sAE |
| IL-8 | ↑↑ | ↑↑ in Ab | n.s. | - | ↑↑ in NORSE | ↑↑ in NORSE | - |
| IL-9 | ↑↑ | n.s. | n.s. | - | - | - | - |
| IL-10 | ↑↑ | n.s. | n.s. | ↑↑ in GADA+ | n.s. | ↑↑ in NORSE | n.s. |
| IL-15 | ↑↑ | n.s. | n.s. | - | - | - | - |
| IL-20 | ↑↑ | - | - | - | - | - | - |
| VEGF-A | ↑↑ | n.s. | ↑↑ in Ab- | - | n.s. | n.s. | - |
| TNF-b | ↑↑ | ↑↑ in Ab | n.s. | - | - | - | - |
| LIF | ↑↑ | - | - | - | - | - | - |
| TSLP | ↑↑ | - | - | - | - | - | - |
| EGF | ↑ | n.s. | n.s. | - | - | - | - |
| MCP-3 (CCL7) | ↑ | n.s. | n.s. | - | - | - | - |
| IL-1A | ↑ | n.s. | n.s. | - | - | - | - |
| IL-13 | ↑ | n.s. | ↑↑ in Ab+ | - | - | - | - |
| IL-16 | ↑ | - | - | - | - | - | - |
| IL-23 | ↑ | - | - | - | - | - | - |
| IL-28A | ↑ | - | - | - | - | - | - |
| TNF-a | n.s. | n.s. | n.s. | - | n.s. | ↑↑ in NORSE | - |
| IL-1B | n.s. | n.s. | n.s. | - | n.s. | n.s. | - |
| IL-12 | n.s. | ↑↑ in Ab | n.s. | - | n.s. | ↑↑ in NORSE | - |
| IL-17A | n.s. | n.s. | n.s. | - | n.s. | n.s. | ↑↑ in sAE |
| RANTES | n.s. | n.s. | ↑↑ in Ab+ | - | - | - | - |
| SDF-1b (CXCL12) | n.s. | - | - | - | n.s. | ↑↑ in NORSE | ↑↑ in sAE |
| MCP-1 (CCL2) | n.s. | n.s. | n.s. | - | ↑↑ in NORSE | ↑↑ in NORSE | - |
| **HMGB1** | - | - | - | - | n.s. | ↑↑ in NORSE | ↑↑ in sAE |
| **INFg** | - | ↑↑ in Ab | n.s. | - | - | - | - |
| **bNGF** | - | ↑↑ in Ab | n.s. | - | - | - | - |

Legend: ↑↑: significant increase, ↑: notable trending increase, n.s.: no significant change reported, -: not measured, Ab+: antibody positive autoimmune epilepsy, Ab-: antibody negative autoimmune epilepsy, sAE: suspected autoimmune epilepsy, GADA+: glutamic acid decarboxylase antibodies positive.

In our study, we did not detect significant changes in the levels of IL-12, IL-17A, or SDF-1b/CXCL12, diverging from prior reports^11,16,17,19,20^. The absence of significant differences of these cytokine levels may be attributable to differences between cohort characteristics, including size and participant composition. For example, Gillinder et al. compared spAAE patients to refractory, non-immune mediated epilepsy controls, while our study examined ASM-responsive controls for clinically suspected seronegative AAE patients^19^. Of note, high mobility group box 1 protein (HMBGB1) was not assessed by our cytokine multiplex assay. Despite these discrepancies, it is important to emphasize that the body of research, including our study, corroborates cytokine perturbations in AAE patients. Furthermore, as they align well with existing literature, our results suggest that IL-6, IL-8, IL-10, IL-13, VEGF-A, and TNF-b could be putative cytokine biomarkers for snAAE^16,17,19^, and may be of use in clinical evaluations of snAAE patients.

Among the snAAE significant cytokines, GM-CSF, MCP-2/CCL8, MIP-1a/CCL3, IL-1RA, IL-9, IL-15, IL-16, IL-20, IL-28A, LIF, and TSLP were novel biomarkers that we identified for the first time in snAAE patients. Notably GM-CSF is implicated in increasing vascular permeability as well as modulating the blood-brain barrier and immune cell migration into the CNS. This cytokine has also been linked to various neurological autoimmune conditions in prior studies^21,22^. Furthermore, MCP-3 (CCL7), a leukocyte chemoattractant, plays a role in recruiting monocytes, granulocytes, dendritic cells, natural killer cells, and activated T cells^23^, likely working in conjunction with GM-CSF and VEGF-A to increase immune infiltration into the CNS.

Interestingly, the PhIP-seq analysis of ∼24,000 analytes failed to identify autoantibodies targeting neuronal channel proteins in snAAE participants, which have been previously implicated in spAAE pathophysiology^24,25^. This finding suggests that simply expanding the scope of autoantibody screens in the snAAE patient population may be inadequate to enhance diagnostic power. Rather, based on our results, we propose that cytokine analysis represents a more promising avenue for identifying immune dysregulation in snAAE.

Beyond serving as a diagnostic aid, cytokine levels may also provide a useful metric in quantifying epilepsy disease severity and/or disease activity, as is done in other conditions including sepsis, heart disease, and rheumatoid arthritis (RA)^26–28^, particularly in cases where patients are unable to accurately report seizure frequency. The benefits of cytokine quantifications are further enhanced by the affordability and emerging accessibility of testing for many of the identified cytokines in clinical settings.

Notably, snAAE patients’ lack of CNS plasma membrane autoantibodies, including those from our exploratory PhIP-seq analysis, combined with their unique cytokine profile, suggests that the etiology of snAAE may have a greater autoinflammatory component than previously thought. PhIP-seq analysis also demonstrated elevations in the proportion of autoantibodies targeting brain cell membrane proteins in patients with non-immune mediated, ASM-responsive epilepsy. Although the precise reason for the elevated levels of autoantibodies against brain cell membrane proteins is unclear, we hypothesize that that there may be localized inflammation which may contribute to bystander activation and subsequent immune hyperactivity/dysfunction in ASM-responsive epilepsy. Thus, despite well controlled seizure activity, ASM-responsive epilepsy patients may experience disease progression independent of seizure activity (PISA), similar to progression independent of relapse activity (PIRA) seen in MS^29,30^, coinciding with increases in autoreactivity. The mechanisms underlying these heightened autoantibody levels and the potential role of the immune system in non-AAE patients warrants further investigation.

Our study had several limitations. First, our cohort was of a limited sample size, and thus may have been underpowered to ascertain significance in the weaker, but meaningful differences in cytokines (such as in EGF, MCP-3/CCL7, IL-1A, IL-13, IL-16, and IL-28A). Second, results from cytokine multiplexing and PhIP-seq in the serum may not directly translate to the CSF compartment. Nonetheless, other established neuroimmune biomarkers are more diagnostically accurate in the serum (e.g. AQP4 autoantibodies in neuromyelitis optica spectrum disorder^31^), and thus future studies profiling cytokine levels within both the CSF and serum of AAE patients may be beneficial in clarifying whether there are differences between these compartments in AAE patients. Finally, although our PhIP-seq analysis identified few autoantibodies of interest, this technique only screens for peptides with linear (as opposed to conformational) epitopes^32^. This limitation is especially relevant when considering that a significant percentage of autoantibodies in humans target conformational epitopes rather than their linear counterparts and, further, many antibodies against channels specifically target channel complexes^33^. Despite these limitations, the results of this study demonstrate notable cytokine differences in snAAE patients, and provide a strong basis for the possible inclusion of cytokines in AAE diagnosis.

Our study constitutes the most extensive panel of assayed cytokines and autoantibodies in a direct comparison of patients with snAAE to ASM-responsive epilepsy, healthy controls, or patients with other neuroinflammatory diseases (MS). While we did not find clinically significant, neuronal specific autoantibodies in snAAE, we identified a specific cytokine signature of IL-6, IL-8, IL-10, IL-13, VEGF-A, and TNF-b, showing the most diagnostic promise in snAAE, as these cytokine perturbations have been replicated across multiple studies. In addition, we demonstrate GM-CSF, MCP-2/CCL8, MIP-1a/CCL3, IL-1RA, IL-9, IL-15, IL-16, IL-20, IL-28A, LIF, and TSLP elevations as novel cytokine biomarkers for snAAE. These findings substantiate the presence of immune dysregulation in a subset of individuals with refractory epilepsy and emphasize the necessity for a broader immune workup in the diagnosis of AAE, including autoantibody and cytokine profiling early in the disease course. Lastly, we believe that our results provide important clinical implications for the care of epilepsy patients. By broadening the immune assessment in patients with refractory epilepsy, it may be possible to identify spAAE and snAAE earlier, towards reducing the morbidity and mortality of individuals living with refractory epilepsy.

## Acknowledgements

We are grateful to the participants of this study. We are also thankful for the administrative support from the Dell Medical School Neurology Department, including Sage Shaw.

## References

1. French JA. Refractory epilepsy: clinical overview. Epilepsia. 2007; 48 Suppl 1:3–7.

2. Husari KS, Dubey D. Autoimmune Epilepsy. Neurotherapeutics. 2019; 16(3):685–702.

3. Berger B, Hauck S, Runge K, Tebartz van Elst L, Rauer S, Endres D. Therapy response in seronegative versus seropositive autoimmune encephalitis. Front Immunol. 2023; 14:1196110.

4. Geis C, Planagumà J, Carreño M, Graus F, Dalmau J. Autoimmune seizures and epilepsy. J Clin Invest. 2019; 129(3):926–40.

5. Dubey D, Pittock SJ, McKeon A. Antibody Prevalence in Epilepsy and Encephalopathy score: Increased specificity and applicability. Epilepsia. 2019; 60(2):367–9.

6. Dubey D, Kothapalli N, McKeon A, Flanagan EP, Lennon VA, Klein CJ, et al. Predictors of neural-specific autoantibodies and immunotherapy response in patients with cognitive dysfunction. J Neuroimmunol. 2018; 323:62–72.

7. Mikaeloff Y, Jambaqué I, Hertz-Pannier L, Zamfirescu A, Adamsbaum C, Plouin P, et al. Devastating epileptic encephalopathy in school-aged children (DESC): a pseudo encephalitis. Epilepsy Res. 2006; 69(1):67–79.

8. Granata T, Andermann F. Rasmussen encephalitis. Handb Clin Neurol. 2013; 111:511–9.

9. Nickels K. NORSE Versus FIRES: What’s in a Name? Epilepsy Curr. 2018; 18(5):301–3.

10. Steriade C, Gillinder L, Rickett K, Hartel G, Higdon L, Britton J, et al. Discerning the Role of Autoimmunity and Autoantibodies in Epilepsy: A Review. JAMA Neurol. 2021; 78(11):1383–90.

11. Han Y, Yang L, Liu X, Feng Y, Pang Z, Lin Y. HMGB1/CXCL12-Mediated Immunity and Th17 Cells Might Underlie Highly Suspected Autoimmune Epilepsy in Elderly Individuals. Neuropsychiatr Dis Treat. 2020; 16:1285–93.

12. Mohan D, Wansley DL, Sie BM, Noon MS, Baer AN, Laserson U, et al. PhIP-Seq characterization of serum antibodies using oligonucleotide-encoded peptidomes. Nat Protoc. 2018; 13(9):1958–78.

13. Uhlén M, Fagerberg L, Hallström BM, Lindskog C, Oksvold P, Mardinoglu A, et al. Proteomics. Tissue-based map of the human proteome. Science. 2015; 347(6220):1260419.

14. Seal RL, Braschi B, Gray K, Jones TEM, Tweedie S, Haim-Vilmovsky L, et al. Genenames.org: the HGNC resources in 2023. Nucleic Acids Res. 2023; 51(D1):D1003–9.

15. Marcuzzi A, Melloni E, Zauli G, Romani A, Secchiero P, Maximova N, et al. Autoinflammatory Diseases and Cytokine Storms-Imbalances of Innate and Adaptative Immunity. Int J Mol Sci. 2021; 22(20):11241.

16. Basnyat P, Peltola M, Raitanen J, Liimatainen S, Rainesalo S, Pesu M, et al. Elevated IL-6 plasma levels are associated with GAD antibodies-associated autoimmune epilepsy. Front Cell Neurosci. 2023; 17:1129907.

17. Billiau AD, Witters P, Ceulemans B, Kasran A, Wouters C, Lagae L. Intravenous immunoglobulins in refractory childhood-onset epilepsy: effects on seizure frequency, EEG activity, and cerebrospinal fluid cytokine profile. Epilepsia. 2007; 48(9):1739–49.

18. Hanin A, Cespedes J, Dorgham K, Pulluru Y, Gopaul M, Gorochov G, et al. Cytokines in New-Onset Refractory Status Epilepticus Predict Outcomes. Ann Neurol. 2023; 94(1):75–90.

19. Gillinder L, McCombe P, Powell T, Hartel G, Gillis D, Rojas IL, et al. Cytokines as a marker of central nervous system autoantibody associated epilepsy. Epilepsy Res. 2021; 176:106708.

20. Cristina de Brito Toscano E, Leandro Marciano Vieira É, Boni Rocha Dias B, Vidigal Caliari M, Paula Gonçalves A, Varela Giannetti A, et al. NLRP3 and NLRP1 inflammasomes are up-regulated in patients with mesial temporal lobe epilepsy and may contribute to overexpression of caspase-1 and IL-β in sclerotic hippocampi. Brain Res. 2021; 1752:147230.

21. Lotfi N, Thome R, Rezaei N, Zhang G-X, Rezaei A, Rostami A, et al. Roles of GM-CSF in the Pathogenesis of Autoimmune Diseases: An Update. Front Immunol. 2019; 10:1265.

22. Carvalho JF, Blank M, Shoenfeld Y. Vascular endothelial growth factor (VEGF) in autoimmune diseases. J Clin Immunol. 2007; 27(3):246–56.

23. Menten P, Wuyts A, Van Damme J. Monocyte chemotactic protein-3. Eur Cytokine Netw. 2001; 12(4):554–60.

24. Toledano M, Pittock SJ. Autoimmune Epilepsy. Semin Neurol. 2015; 35(3):245–58.

25. Bien CG, Holtkamp M. “Autoimmune Epilepsy”: Encephalitis With Autoantibodies for Epileptologists. Epilepsy Curr. 2017; 17(3):134–41.

26. Bozza FA, Salluh JI, Japiassu AM, Soares M, Assis EF, Gomes RN, et al. Cytokine profiles as markers of disease severity in sepsis: a multiplex analysis. Crit Care. 2007; 11(2):R49.

27. Guilherme L, Cury P, Demarchi LMF, Coelho V, Abel L, Lopez AP, et al. Rheumatic heart disease: proinflammatory cytokines play a role in the progression and maintenance of valvular lesions. Am J Pathol. 2004; 165(5):1583–91.

28. McInnes IB, Schett G. Cytokines in the pathogenesis of rheumatoid arthritis. Nat Rev Immunol. 2007; 7(6):429–42.

29. Sharrad D, Chugh P, Slee M, Bacchi S. Defining progression independent of relapse activity (PIRA) in adult patients with relapsing multiple sclerosis: A systematic review✰. Mult Scler Relat Disord. 2023; 78:104899.

30. Kappos L, Wolinsky JS, Giovannoni G, Arnold DL, Wang Q, Bernasconi C, et al. Contribution of Relapse-Independent Progression vs Relapse-Associated Worsening to Overall Confirmed Disability Accumulation in Typical Relapsing Multiple Sclerosis in a Pooled Analysis of 2 Randomized Clinical Trials. JAMA Neurol. 2020; 77(9):1132–40.

31. Majed M, Fryer JP, McKeon A, Lennon VA, Pittock SJ. Clinical utility of testing AQP4-IgG in CSF: Guidance for physicians. Neurol Neuroimmunol Neuroinflamm. 2016; 3(3):e231.

32. Tiu CK, Zhu F, Wang L-F, de Alwis R. Phage ImmunoPrecipitation Sequencing (PhIP-Seq): The Promise of High Throughput Serology. Pathogens. 2022; 11(5):568.

33. DeMarshall C, Sarkar A, Nagele EP, Goldwaser E, Godsey G, Acharya NK, et al. Utility of autoantibodies as biomarkers for diagnosis and staging of neurodegenerative diseases. Int Rev Neurobiol. 2015; 122:1–51.

