## Supplemental Figure 1 for "Altered Cytokine Profile in Clinically Suspected Seronegative Autoimmune Associated Epilepsy"

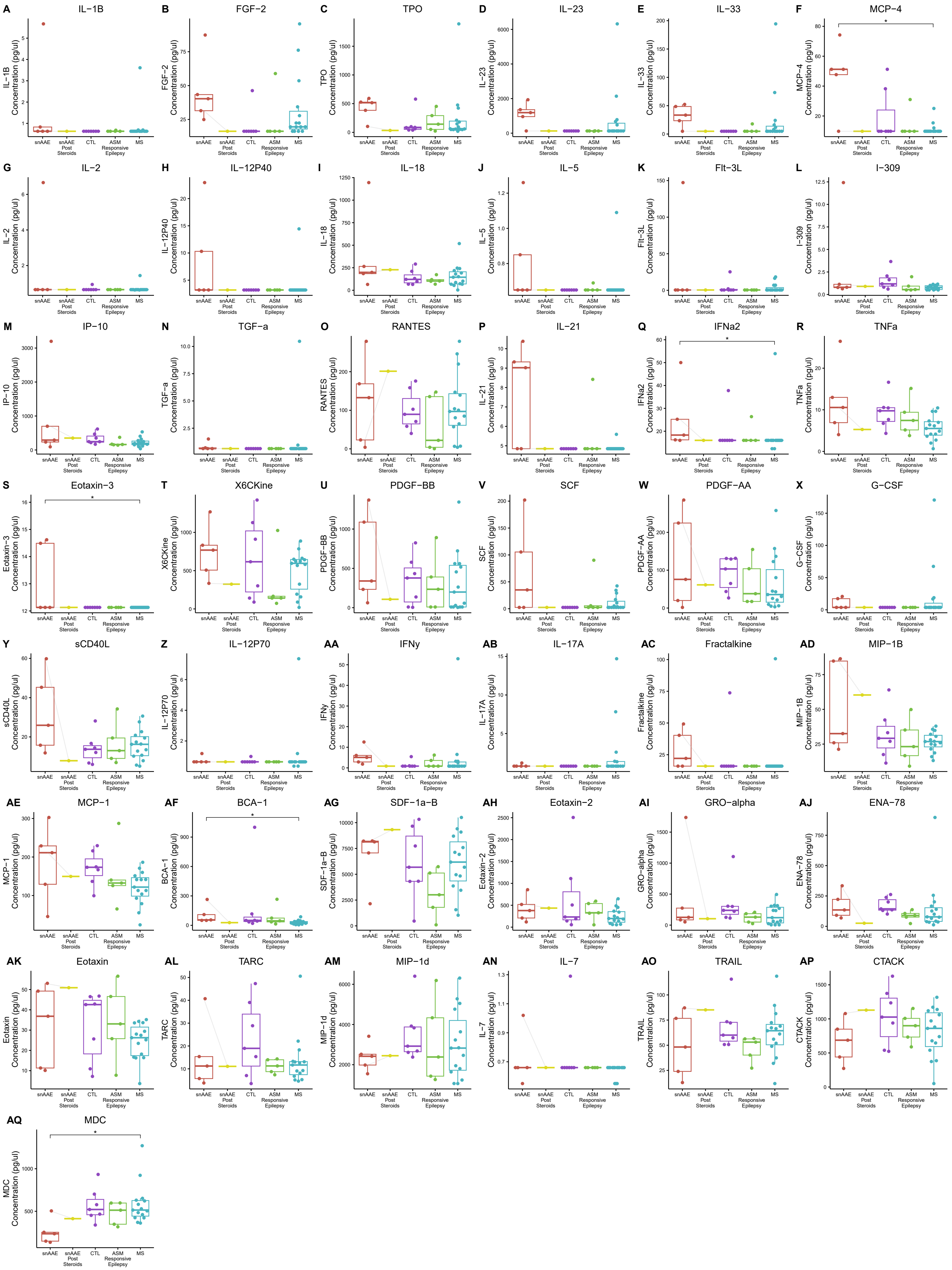


**Supplemental Figure 1 –** Boxplots of the remaining cytokines from the multiplex assay (associated with Figure 1).
